# Highlighting the importance of semantics in COVID-19 fake news detection using a CNN hashmap color-based method

**DOI:** 10.1101/2022.07.24.22277975

**Authors:** Maryam El Azhari

## Abstract

With the exponential development and exploitation of social media sites and platforms such as facebook, twitter and instagram, a diversity type of news are reached to the users,resulting in a major influence on human health and safety.Spreading misinformation and disinformation during the Covid-19 pandemic has become increasingly significant. Although it is usually not a criminal act, it can cause serious endangerment to public health. Such infodemic movement is often lead to advance geopolitical interests by the states, to achieve some sort of profit by some opportunists and individuals or discredit official sources. Hence,it has become crucial to automate the detection of fake news in order to shield people from any harmful repercussions. In this paper, the importance of semantics in Covid-19 fake news detection is highlighted based on a convolutional neural network classifier and a hashmap color-based technique. The experiments are performed with CoAID(Covid-19 heAlthcare mIsinformation Dataset),and the results prove that the loss of semantics yields to a poor performance of the classifier. This implicates additional constraints to the training images,with focus on creating a CNN-based color hashmap classifier that includes anterior and posterior neighbors.

## 1 Introduction

On March 11, 2020, Coronavirus disease (COVID-19) caused by the new coronavirus (SARS) CoV-2) was declared a pandemic by World Health Organization (WHO). The coronavirus pandemic has drastically influenced the world economy and caused death of millions people around the world [1]. As the COVID-19 disease has become widely spread across the world,some social disruptions have accompanied this rapid downturn due to misinformation.As a matter of fact,many fake cures have claimed fraudulent products to cure the coronavirus thus causing potential threats to people’s lives. A study published in the American Journal of Tropical Medicine and Hygiene estimates that nearly 6000 people entered hospitals as a result of unproven medical products to cure the disease whilst other people were announced dead after drinking methanol or alcohol-based cleaning solutions [2]. Facebook and Instagram (owned by Meta) have claimed that 20m pieces of harmful misinformation had been removed.Moreover, nearly 3,000 accounts,groups and pages have been banned for repeatedly violating the rules. Although many social media users who spread fake news over the internet are real people,there is still a vast majority of fake news contributors, that fall into three main categories: Cyborgs,Trolls and Bots [3]. A bot generate content and interact with internet users in an automatic manner.They are often referred to as automated social accounts programmed to spread content quickly,sometimes they are harmless but other times they can be programmed to mimic humans in an effort to hide their motives. Bots are malicious contributors to fake news only if they are programmed for that purpose. Trolls are internet users who post content intended to anger,irritate or annoy. They may use a fake name or profile picture, and they are often just people looking to start up trouble. Cyborgs are type of bots occasionally run by an actual human, usually as a way to make a bot appear more like a person. The following five categories of fake news detection approaches were discussed in more detail in [4–10]: language approach,topic-agnostic approach,machine learning approach,knowledge-based approach and hybrid approach. In this paper,the importance of semantics for fake news detection is studied in detail, using a convolutional neural network classifier and a hashmap color-based technique. The reminder of this paper is organized as follows: Section 2 introduces the proposed fake news detection paradigm. Section 3 presents the experiments and analyzes the results being found. And finally, Section 4 concludes this paper.

## 2 A new fake news detection paradigm

Detecting fake news has become one of the most important tasks to be accomplished by artificial intelligence researchers. Machine learning and Deep Learning, are the two most commonly used approaches for detection. Before proceeding with applying a CNN hashmap color-based classification method,text prepossessing is executed on the available dataset to generate two non intersected word clouds, where each word cloud represents a class type of the dataset. The main idea behind this approach is to reduce the computational complexity of processing an image,but also to create a fingerprinted image of hashed colors that can be classified into fake or real news category. Figure 1 shows the procedure of text cleaning before creating the non-intersected word clouds. This procedure is applicable for each entry of the dataset.

**Fig. 1:**
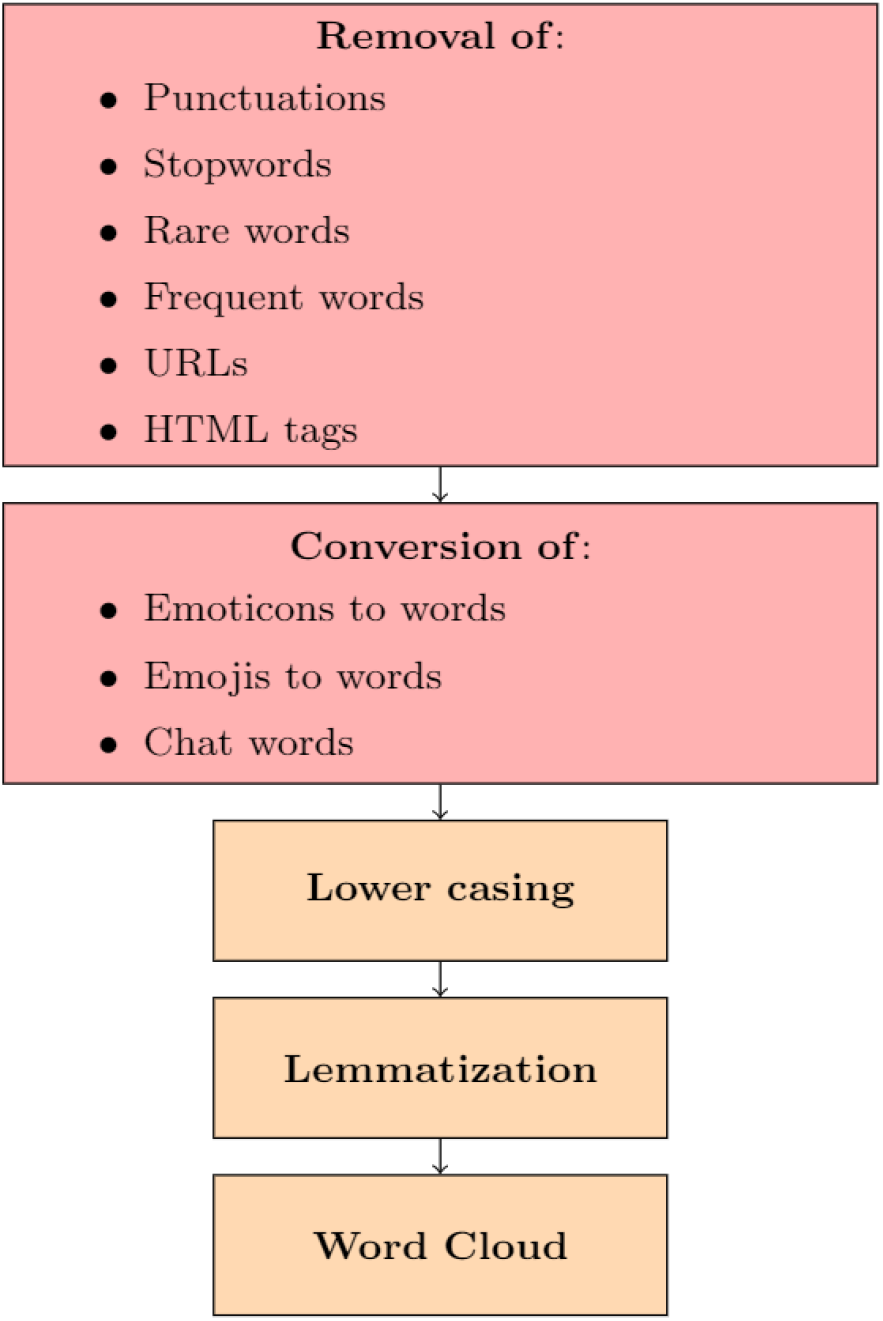
Text preprocessing.

Let *WC* = ⋃_*i*∈*I*_*WC*_*f i*_ ⋃ ⋃_*j*∈*J*_ *WC*_*rj*_ where I and J refer to the number of fake news and real news dataset entries respectively. After obtaining the words cloud set *WC*,the proximity matrices for *WC*_*f*_ and *WC*_*rj*_ subsets are calculated.Each entry of the matrix holds the probabilities that a *word A* exits in the vicinity of *word B* within a range of 1. Let *W* be a random word that belongs to *WC*_*kc*_ where *k ∈ f, r* and *c I*, ∈ *J*. The proximity matrix is defined as follows:

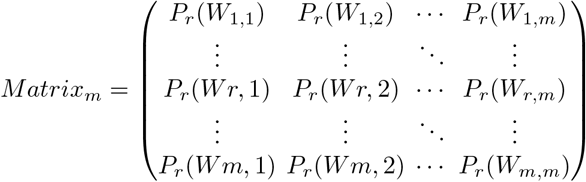

where:

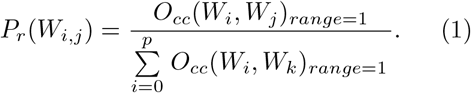

The proximity matrix can be written as :

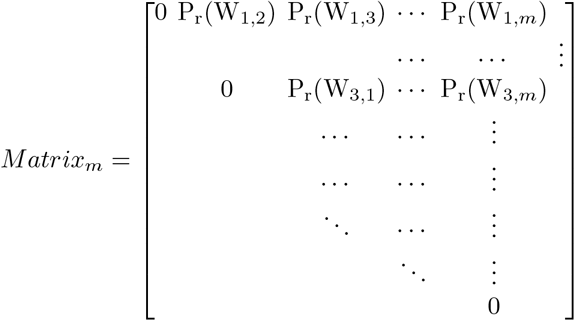

where 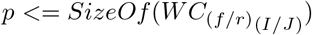 and *k* ≠ *j*

For a vicinity range *>*= 2,*P*_*r*_ formula is represented as follows:

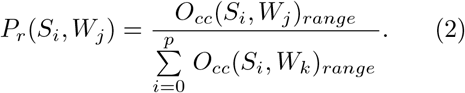

where *S* = {*W*_*i*_, *W*_2_, &, *W*_*d*_}, *d <*= *m* and *range* = *SizeOf* (*S*_*i*_). After defining the proximity matrix, a 1D data clustering is performed on each *Row*_*i*_ of *Matrix*_*m*_ to create clusters of probabilities. Each cluster is assigned a priority to be selected with respect to the probability value, and all the words belonging to a cluster inherit the same priority.Clusters with a high priority are the first to be selected to fill in the color map images.

### 2.1 The theoretical approach of creating a color map images

Creating color map images consists in giving a unique representation of a word *W* ∈ *WC*_*f/r*_ by mapping each word to a unique color.To this end,the md5 hash of each word can be used to generate the R, G and B values by using the first three bytes of the md5 hash,however,the resulting color can turn faint due to the randomness of R,G and B components.To resolve this issue,the color is first generated in HSV color space with regulated saturation and brightness,followed by conversion to RGB model.The colorsys module is used to define the bidirectional conversions of color values between colors expressed in the RGB (Red Green Blue) and HSV (Hue Saturation Value). Figure 2 shows an example of Word Clouds 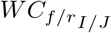 and Word Clouds *HSV_RGB*(*W*_*i*_).

**Fig. 2:**
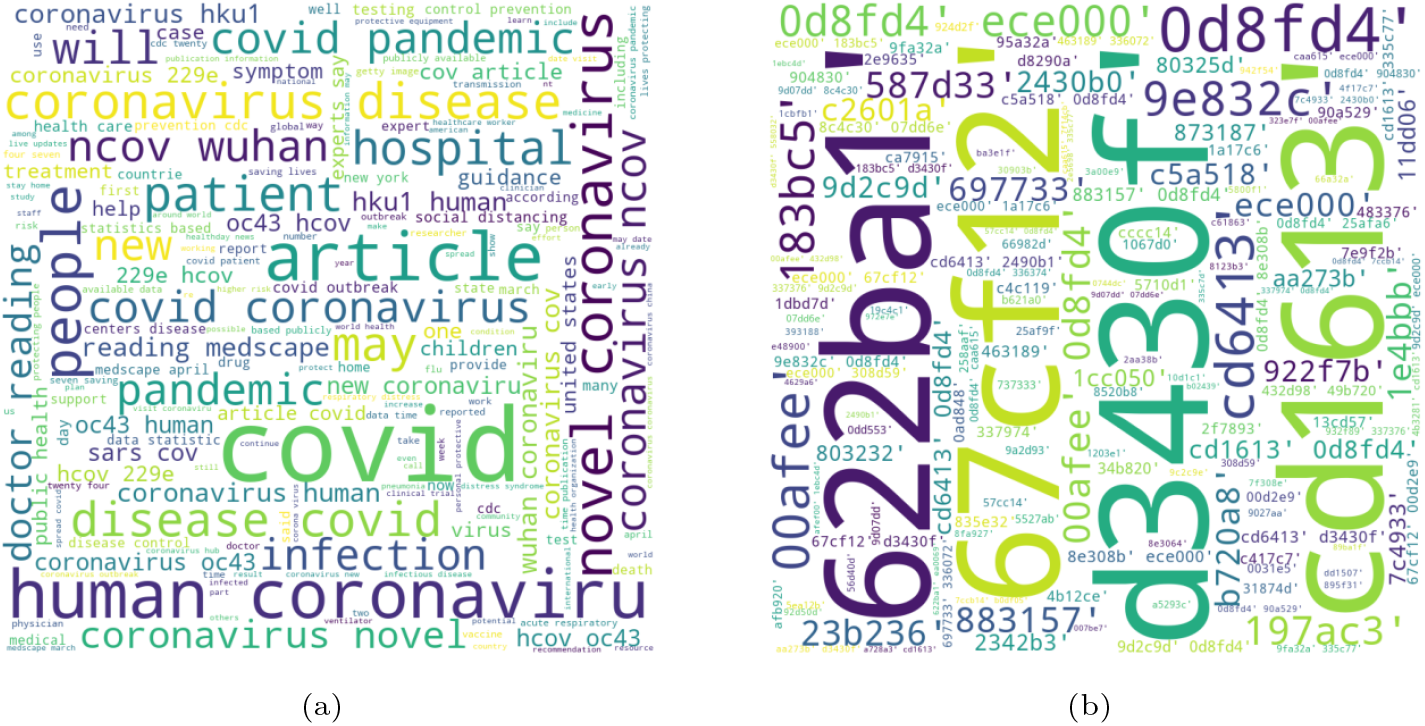
(a) Word Clouds 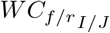 (b) Word Clouds *HSV_RGB*(*W*_*i*_)

The distribution of RGB hash mapping colors in an image is constrained by the proximity Matrix.A particular color occupies one or more pixels within an image where the dimensions are initially defined by the length of words cloud subset *WC*_*f/r*_. A dimension thresh-hold is defined for word clouds subsets that exceed the size limit.The coloring process starts with choosing a random word *W*_*i*_ *i*.*e* a random *row*_*i*_ from the proximity matrix.*N* number of pixels values are set to *HSV_RGB*(*W*_*i*_) starting from index (0, 0) (Where *N* ≥ 1). Setting the *N* pixels value of neighbors within 1 range is done in a clockwise direction,where the corresponding *HSV_RGB*(*W*_*j*_) value is selected randomly from a *cluster*_*i*_ which belongs to the list of clusters *Clusters*(*W*_*i*_) with the highest priority.The random selection is performed on a list of words (*i*.*eHSV_RGB*(*W*)) having the maximum *Counter* value. The *Counter* of a word *W*_*i*_((*i*.*eHSV_RGB*(*W*_*i*_))) keeps track of its usage and is decremented each time it is selected.The *Counter* of a word *W*_*j*_ is defined as below:

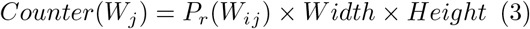

where *Counter*(*W*_*j*_) ∈ ℕ

Once the value of *Counter*(*W*_*j*_) reaches zero,a new *HSV_RGB*(*W*_*j*_) is selected among the same cluster having the maximum *Counter*(*W*_*j*_) value,if the cluster set *Cluster*(*W*_*i*_) is void then the cluster ranked in second place is selected and the procedure is repeated until the entire clusters are circulated. *Counter*(*W*_*j*_) is reset to its initial value and the same process is repeated for other rounds.The random selection of *Row*_*i*_ from *Matrix*_*m*_ and *HSV_RGB*(*W*_*j*_) from *Cluster*(*W*_*i*_) generate more images for training alongside with Random Image Generator. The resulting image is represented as follows:

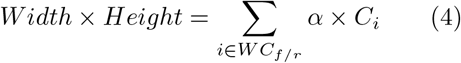

where :

- *C*_*i*_ is the RGB representation of *W*_*i*_
- *Width* ≤ *Max*(*SizeOf* (WC_*f/r*_))
- *Height* ≤ *Max*(*SizeOf* (WC_*f/r*_))
- *Width* ×*Height* ≤Dim_*thresh*_
- 0 ≤*α <*Max(SizeOf(*WC*_*f/r*_))

## 3 Experiments and results

In this paper,the performance of the proposed method is studied based upon the presumption that all the words in *WC* have the same probability to be located within the 1-range of a word *W*_*i*_. This hypothesis allows conducting a partial study of the proposed method by investigating the impact of semantics when performing infodemic classification. Experiments have been performed using CoAID [11].CoAID includes a diversity of fake news on websites and social platforms along with related user engagements. CoAID includes 1,896 news, 183,564 related user engagements, 516 social platform posts about COVID-19, and ground truth labels. The dataset is available at: https://github.com/cuilimeng/CoAID. A convolution neural network(CNN) is implemented to perform image classification.As can be shown in Figure 3,the CNN model focuses on having 5 essential convolution layers of 32,64 and 128 filters with 3 × 3 kernels,a stride of 2 and same padding.The CNN model ends with 2 fully connected layers and a sigmoid for output.The sequential method is used to create a sequential model.After initializing the model,the following layers are embedded sequentially posterior to the convolution layers:an activation layer with Relu function,a batch normalization layer,a maxpool layer of 3×3 pool size,a stride of 2×2 and a dropout layer.After creating all the convolution layers,the data is flattened and passed to a dense layer with 1024 neurons followed by a dense Softmax layer with 2 units. The images are augmented to create a diversity of input data and provide a balanced class distribution. The augmented images are resized to (224 × 224 × 3) and normalized between 0 and 1. The resulting output is randomly split into training,validation and test sets with respect to the mask vector *<* 80, 10, 10 *>*

**Fig. 3:**
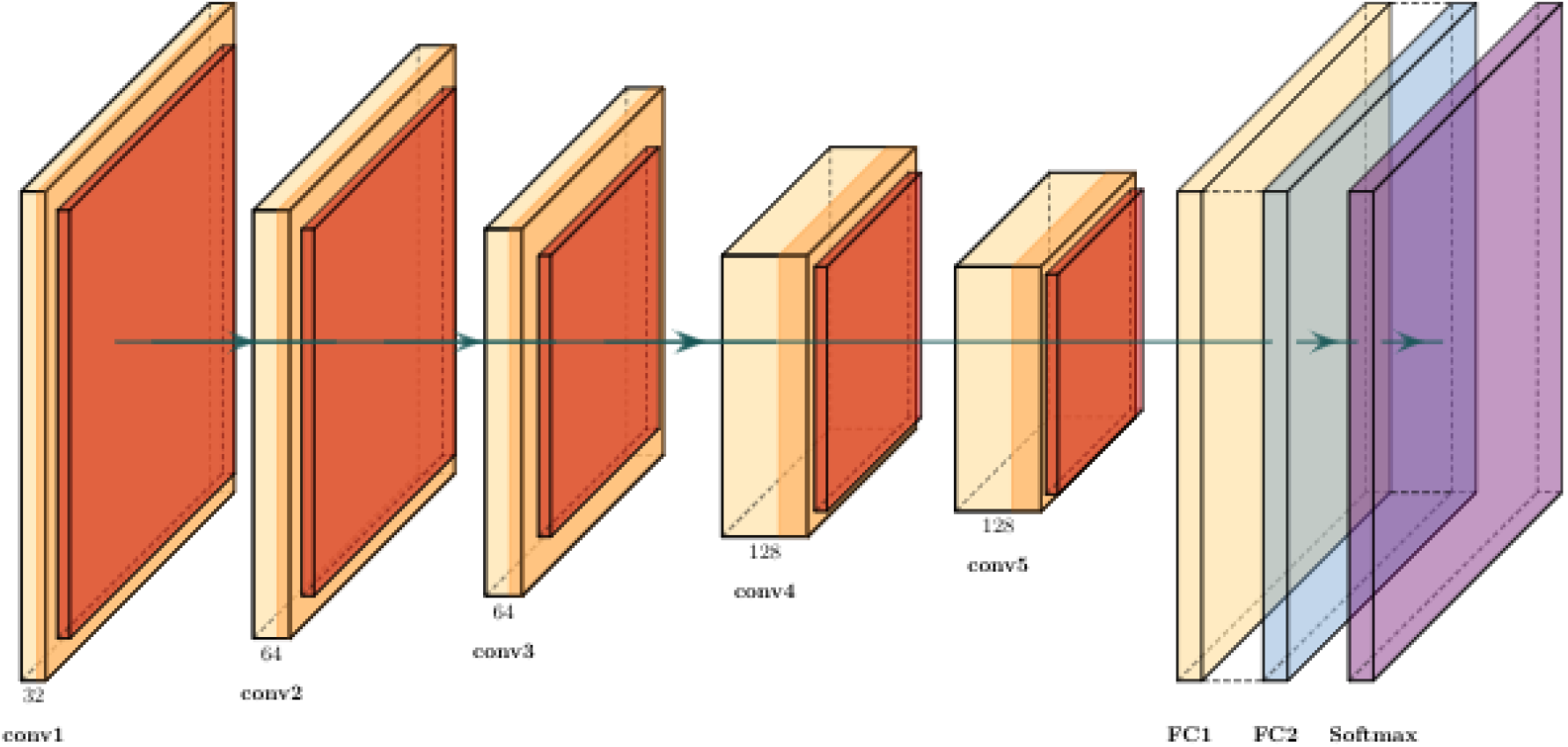
CNN architecture for fake news detection.

There are various evaluation metrics to assess the performance of the deep learning models.All the following metrics used in this study are based on the confusion matrix defined in Figure 4: precision,recall,F1 score and accuracy.

**Fig. 4:**
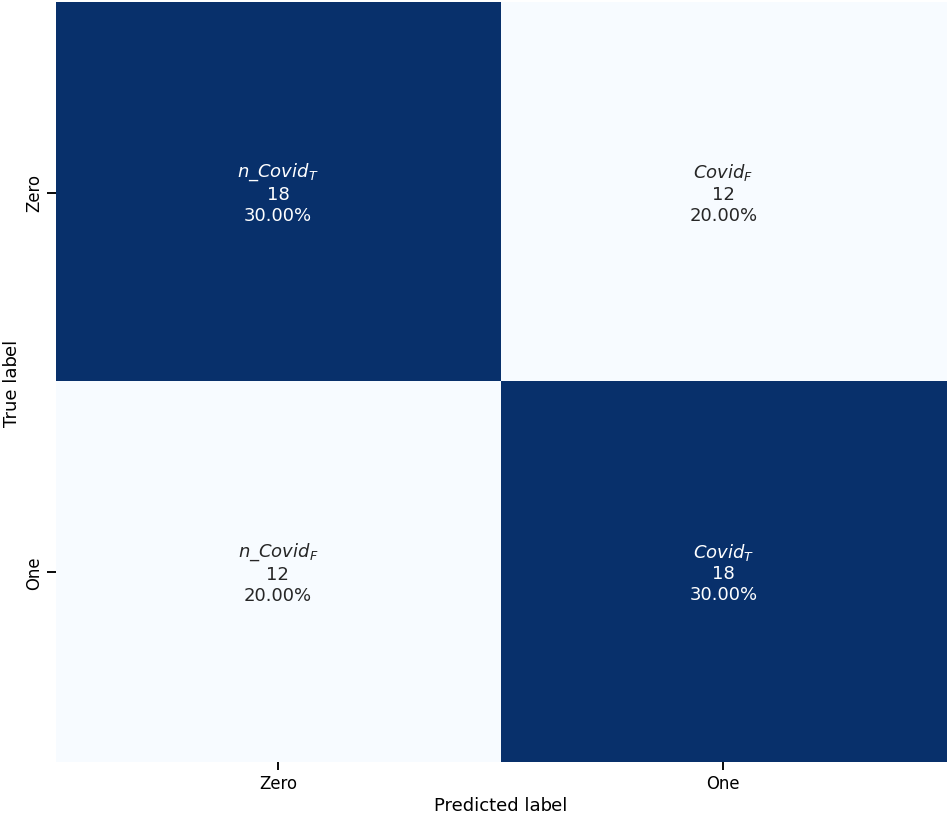
Confusion matrix for *Covid* − 19 detection.

### 3.1 Performance evaluation metrics

Recall is a metric that defines the proportion of actual covid-19 positive cases that were predicted correctly.The definition of recall is given by then formula below:

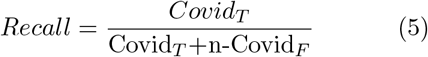

Precision is a metric used to define the proportion of positive predictions that were actually correct.Precision is depicted in the formula below:

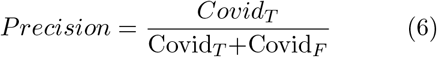

F1-score metric measures the quality of binary classification problems.It is often used to select a model based on a balance between recall and precision.The F1-score metric is defined as:

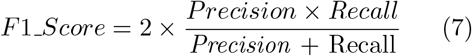

The accuracy metric measures how often the model predicts correctly.The accuracy formula is defined as:

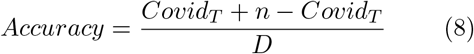

where:

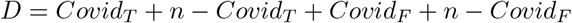

### 3.2 Results and discussion

The results from Table 1 demonstrate a poor performance of the proposed CNN classifier when semantics is ignored.All the aforementioned evaluation metrics scored equally with a percentage of 60%.When a word belonging to the same subset is chosen randomly to construct a hash mapped color image,the information that holds the truthfulness of certain news is lost.This information is embodied in the proximity matrix defined in section 2 with equation 2. The proximity matrix presented in this paper is equivalent to context in word embedding [12] where words are represented with vectors, and the context of a word is captured to find out similarity with other words. A semantic-based CNN classifier will consider a onestep anterior and posterior neighbors of a certain word to construct a hash mapped color image. In fact,a typical word structure within a text implies 8 neighbors for each word *W*_*i*_ located at least one-step away from all edges,where *W*_*i*_ is the center cell of a cube with side length of 3 cells.When all 8 neighbors are considered for building the CNN classifier,new constraints related to text format are imposed,therefore the classifier will be over-fitting on the text data.To make a more generalized CNN model,only the one-step neighbors that are horizontally aligned with a word *W*_*i*_ (i.e. anterior and posterior neighbors) are nominated for use, to create a hash mapped color image.

**Table 1:**
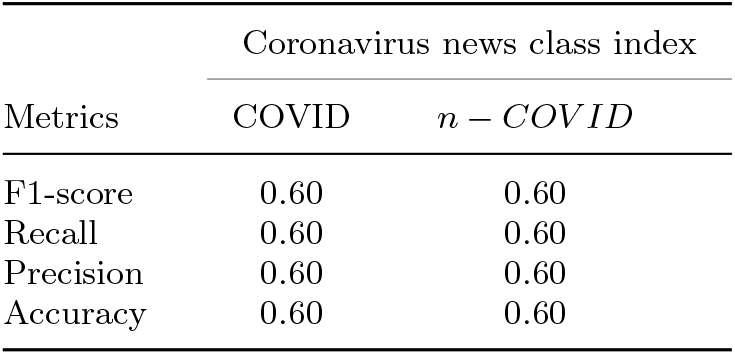
Experimental results

| Metrics | Coronavirus news class index |  |
| --- | --- | --- |
| | COVID | $n - COVID$ |
| F1-score | 0.60 | 0.60 |
| Recall | 0.60 | 0.60 |
| Precision | 0.60 | 0.60 |
| Accuracy | 0.60 | 0.60 |

The following commonly used models are further investigated to highlight the impact of semantic on fake news detection: Bag-Of-Words(BOW),Term Frequency Inverse Document Frequency(TF-IDF), Long Short-Term Memory (LSTM) and n-gram.

#### BOW

The BOW model is based on representing the dataset as a bag of words vector. In this model, a vocabulary of unique words is first built from every subset of the dataset. The occurrence of these words is marked with 1s and 0s to generate word count vectors E.g. [1, 2, 0, 1, 0, 1, 1, 0, 0&]

#### TF-IDF

TF-IDF was initially invented to find results that are most relevant to the documents of interest. TF-IDF is a measure that indicates the number of times a particular word *w* occurs in a document *D* divided by the total number of words in the same document. This measure will help to decide the importance of a word to a given document in a dataset [13].

TF-IDF is calculated by multiplying two metrics:

- The term frequency of a word in a document: this can be defined by a simple count of the word occurrence in a document.
- The inverse document frequency of the word across a set of documents. This refers to how common a word is in the entire dataset. The closer it is to 1,the more rare a word is.

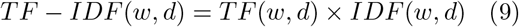

where :

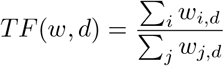

- Σ_*i*_ *w*_*i,d*_ : the number of occurrences of a word *w*_*i*_ in document d.
- Σ_*j*_ *w*_*i,d*_ : the total number of terms *w* in document d.

and :

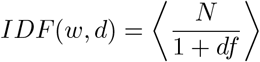

- *N* is the total number of documents.
- *df* is the number of documents with term w.

#### LSTM

LSTM is a recurrent neural network (RNN) architecture that is capable of learning long-term dependencies. The LSTM network has the advantage of remembering information for multiple time intervals through the use of a flow regulator a.k.a. ”gates” [14]. LSTM surpasses the following limits of RNN:

- **Short-term memory**: important information from earlier steps are lost when moving to later ones.
- **Exploding gradient**: this problem occurs when the gradient increases exponentially as the multiplied derivatives get large during back-propagation.
- **Vanishing gradient**: this problem is issued from adding more layers to the neural networks which decreases the gradients of the loss function,and makes the network very hard to train.

#### N-gram

N-gram is a very widely used technique in natural language processing field. It is represented as a sequence of n items from a text data. These items may refer to letters,words or any other subset of words depending on the context of the problem at hand. There are multiple versions of n-gram: 1-gram (unigram),2-gram (bigram),3-gram (trigram),etc. [15]

#### Discussion and Analysis

A random shuffle of CoAID news is performed through an arbitrary number of rounds *Round*_*x*_(*x* is set to 3 in this case of the study),where the model accuracy value is calculated correspondingly to each round. Decision Tree (DT),Gradient Boosting Classifier(GBC),Random Forest(RF),Support Vector Machine (SVM),Logistic Regression (LR),Multinomial Naive Bayes (MNB) and k-nearest neighbors (KNN) algorithms are applied to classify the CoAID news with respect to n-gram methods. Table 2 represents the training and test accuracy results of applying BOW and TF-IDF models to build vocabulary and extract semantics from text data by vectorizing the text sentences. N-gram is considered as a feature, and it is used to maintain the real word order within a sentence. The following study focuses more on investigating the impact of semantics rather than proposing a model that gives the best classification performance. To this end,a total number of 400 dataset entries that combines real and fake news are considered for training and testing the machine learning models. As can be deducted from Table 2, there is an inverse correlation between the training (e.g. test) accuracy and the *n* − *gram* model with respect to BOW and TFIDF. In fact,the longer is the sequence of N words (*bigram < trigram < fourgram < fivegram <* &) the less is the accuracy. When selecting a continuous sequence of N words (*N >* 1),the classification task is constrained by the occurrence of similar sequences in the dataset during the training and test phases. In order to consider the word order within a sequence,it is necessary to have a large dataset to ensure the variety of text sequences,and also to increase the probability of redundant sequence occurrence. This deduction is supported by the results being found in Table 2. In fact,the training accuracy surpasses the test accuracy knowing that the test data represent only 33% of the overall dataset size. Unlike the *n* − *gram* model where *n >* 1,a unigram model will put less constraints on word order and will presumably improve the classification accuracy. In order to investigate further this presumption,a study of the unigram model is conducted over 3 rounds,where the performance of the State-of-the-Art classifiers using BoW and TF-IDF features is evaluated as stated in Table 3. To get a better understanding of the results being found,the average value of the training and test accuracy is calculated using equation 10, and the results are presented in Table 4. As opposed to the sequence of words,a unigram model gives better results in terms of training and test accuracy.For instance,the average training accuracy with a unigram model using BOW feature is equal to 0.94 versus 0.79 for a *n* − *gram* model. Likewise the average test accuracy with a unigram model using *TF* − *IDF* feature is equal to 0.81 versus 0.59 for an *n* − *gram* model. This performance is due to the abundance of data vis-à-vis the unconstrained word order, and it also proves that *BOW* and *TF* − *IDF* models are dependant on word frequency rather that word arrangement. When the word order is shuffled randomly with respect to a unigram model,*BOW* and *TF* − *IDF* show no difference in performance and the results are very approximate over multiple rounds.

**Table 2:** Classification accuracies using Bag-of-words and TF-IDF

| **BOW: Round 1** |  |  |  |  |  |
| --- | --- | --- | --- | --- | --- |
| Bag-of-words (BOW) |  |  |  |  |  |
| Classifier name | Metrics | 2-gram | 3-gram | 4-gram | 5-gram |
| DT | training accuracy | 0.9975 | 0.935 | 0.7725 | 0.6725 |
|  | test accuracy | 0.82 | 0.65 | 0.55 | 0.56 |
| GBC | training accuracy | 0.9525 | 0.8525 | 0.7575 | 0.6725 |
|  | test accuracy | 0.85 | 0.68 | 0.57 | 0.56 |
| RF | training accuracy | 0.9975 | 0.935 | 0.7725 | .6725 |
|  | test accuracy | 0.82 | 0.74 | 0.56 | 0.56 |
| SVM | training accuracy | 0.9975 | 0.9075 | 0.7675 | 0.6725 |
|  | test accuracy | 0.76 | 0.66 | 0.56 | 0.56 |
| LRM | training accuracy | 0.9875 | 0.915 | 0.7675 | 0.6725 |
|  | test accuracy | 0.81 | 0.72 | 0.57 | 0.56 |
| MNB | training accuracy | 0.875 | 0.8 | 0.7475 | 0.6725 |
|  | test accuracy | 0.71 | 0.64 | 0.57 | 0.56 |
| KNN | training accuracy | 0.71 | 0.735 | 0.55 | 0.575 |
|  | test accuracy | 0.56 | 0.53 | 0.46 | 0.56 |

| **BOW: Round 2** |  |  |  |  |  |
| --- | --- | --- | --- | --- | --- |
| Bag-of-words (BOW) |  |  |  |  |  |
| Classifier name | Metrics | 2-gram | 3-gram | 4-gram | 5-gram |
| DT | training accuracy | 0.9975 | 0.935 | 0.7725 | 0.6725 |
|  | test accuracy | 0.85 | 0.63 | 0.55 | 0.56 |
| GBC | training accuracy | 0.9525 | 0.8525 | 0.7575 | 0.6725 |
|  | test accuracy | 0.85 | 0.68 | 0.57 | 0.56 |
| RF | training accuracy | 0.9975 | 0.935 | 0.7725 | 0.6725 |
|  | test accuracy | 0.82 | 0.74 | 0.56 | 0.56 |
| SVM | training accuracy | 0.9975 | 0.9075 | 0.7675 | 0.6725 |
|  | test accuracy | 0.76 | 0.66 | 0.56 | 0.56 |
| LRM | training accuracy | 0.9875 | 0.915 | 0.7675 | 0.6725 |
|  | test accuracy | 0.81 | 0.72 | 0.57 | 0.56 |
| MNB | training accuracy | 0.875 | 0.8 | 0.7475 | 0.6725 |
|  | test accuracy | 0.71 | 0.64 | 0.57 | 0.56 |
| KNN | training accuracy | 0.71 | 0.735 | 0.55 | 0.575 |
|  | test accuracy | 0.56 | 0.53 | 0.46 | 0.56 |

**BOW: Round 3**

| Bag-of-words (BOW) |  |  |  |  |  |
| --- | --- | --- | --- | --- | --- |
| Classifier name | Metrics | 2-gram | 3-gram | 4-gram | 5-gram |
| DT | training accuracy | 0.99757 | 0.935 | 0.7725 | 0.6725 |
|  | test accuracy | 0.8 | 0.63 | 0.55 | 0.56 |
| GBC | training accuracy | 0.9525 | 0.8525 | 0.7575 | 0.6725 |
|  | test accuracy | 0.85 | 0.68 | 0.57 | 0.56 |
| RF | training accuracy | 0.9975 | 0.935 | 0.7725 | 0.6725 |
|  | test accuracy | 0.82 | 0.74 | 0.56 | 0.56 |
| SVM | training accuracy | 0.9975 | 0.9075 | 0.7675 | 0.6725 |
|  | test accuracy | 0.76 | 0.66 | 0.56 | 0.6725 |
| LRM | training accuracy | 0.9875 | 0.915 | 0.7675 | 0.6725 |
|  | test accuracy | 0.81 | 0.72 | 0.57 | 0.56 |
| MNB | training accuracy | 0.875 | 0.8 | 0.7475 | 0.6725 |
|  | test accuracy | 0.71 | 0.64 | 0.57 | 0.56 |
| KNN | training accuracy | 0.71 | 0.735 | 0.55 | 0.575 |
|  | test accuracy | 0.56 | 0.53 | 0.46 | 0.56 |

**TF-IDF: Round 1**

| TF-IDF |  |  |  |  |  |
| --- | --- | --- | --- | --- | --- |
| Classifier name | Metrics | 2-gram | 3-gram | 4-gram | 5-gram |
| DT | training accuracy | 1 | 1 | 1 | 1 |
|  | test accuracy | 0.83 | 0.66 | 0.56 | 0.49 |
| GBC | training accuracy | 0.9925 | 0.9225 | 0.7775 | 0.875 |
|  | test accuracy | 0.86 | 0.69 | 0.56 | 0.56 |
| RF | training accuracy | 1 | 0.9975 | 0.9975 | 0.9975 |
|  | test accuracy | 0.68 | 0.49 | 0.44 | 0.44 |
| SVM | training accuracy | 1 | 0.985 | 0.985 | 0.98 |
|  | test accuracy | 0.88 | 0.69 | 0.63 | 0.61 |
| LRM | training accuracy | 1 | 0.985 | 0.985 | 0.98 |
|  | test accuracy | 0.86 | 0.62 | 0.6 | 0.58 |
| MNB | training accuracy | 0.985 | 0.985 | 0.985 | 0.98 |
|  | test accuracy | 0.74 | 0.68 | 0.63 | 0.61 |
| KNN | training accuracy | 0.4875 | 0.4875 | 0.4875 | 0.4875 |
|  | test accuracy | 0.44 | 0.44 | 0.44 | 0.44 |

**TF-IDF: Round 2**

| TF-IDF |  |  |  |  |  |
| --- | --- | --- | --- | --- | --- |
| Classifier name | Metrics | 2-gram | 3-gram | 4-gram | 5-gram |
| DT | training accuracy | 1 | 1 | 1 | 1 |
|  | test accuracy | 0.79 | 0.66 | 0.57 | 0.49 |
| GBC | training accuracy | 0.9925 | 0.9225 | 0.7775 | 0.875 |
|  | test accuracy | 0.86 | 0.69 | 0.56 | 0.56 |
| RF | training accuracy | 1 | 0.9975 | 0.9975 | 0.9975 |
|  | test accuracy | 0.68 | 0.49 | 0.44 | 0.44 |
| SVM | training accuracy | 1 | 0.985 | 0.985 | 0.98 |
|  | test accuracy | 0.88 | 0.69 | 0.63 | 0.61 |
| LRM | training accuracy | 1 | 0.985 | 0.985 | 0.98 |
|  | test accuracy | 0.86 | 0.62 | 0.6 | 0.58 |
| MNB | training accuracy | 0.985 | 0.985 | 0.985 | 0.98 |
|  | test accuracy | 0.74 | 0.68 | 0.63 | 0.61 |
| KNN | training accuracy | 0.4875 | 0.4875 | 0.4875 | 0.4875 |
|  | test accuracy | 0.44 | 0.44 | 0.44 | 0.44 |

**TF-IDF: Round 3**

| TF-IDF |  |  |  |  |  |
| --- | --- | --- | --- | --- | --- |
| Classifier name | Metrics | 2-gram | 3-gram | 4-gram | 5-gram |
| DT | training accuracy | 1 | 1 | 1 | 1 |
|  | test accuracy | 0.84 | 0.65 | 0.56 | 0.49 |
| GBC | training accuracy | 0.9925 | 0.9225 | 0.7775 | 0.875 |
|  | test accuracy | 0.86 | 0.69 | 0.56 | 0.56 |
| RF | training accuracy | 1 | 0.9975 | 0.9975 | 0.9975 |
|  | test accuracy | 0.68 | 0.49 | 0.44 | 0.44 |
| SVM | training accuracy | 1 | 0.985 | 0.985 | 0.98 |
|  | test accuracy | 0.88 | 0.69 | 0.63 | 0.61 |
| LRM | training accuracy | 1 | 0.985 | 0.985 | 0.98 |
|  | test accuracy | 0.86 | 0.62 | 0.6 | 0.58 |
| MNB | training accuracy | 0.985 | 0.985 | 0.985 | 0.98 |
|  | test accuracy | 0.74 | 0.68 | 0.63 | 0.61 |
| KNN | training accuracy | 0.4875 | 0.4875 | 0.4875 | 0.4875 |
|  | test accuracy | 0.44 | 0.44 | 0.44 | 0.44 |

**Table 3:**
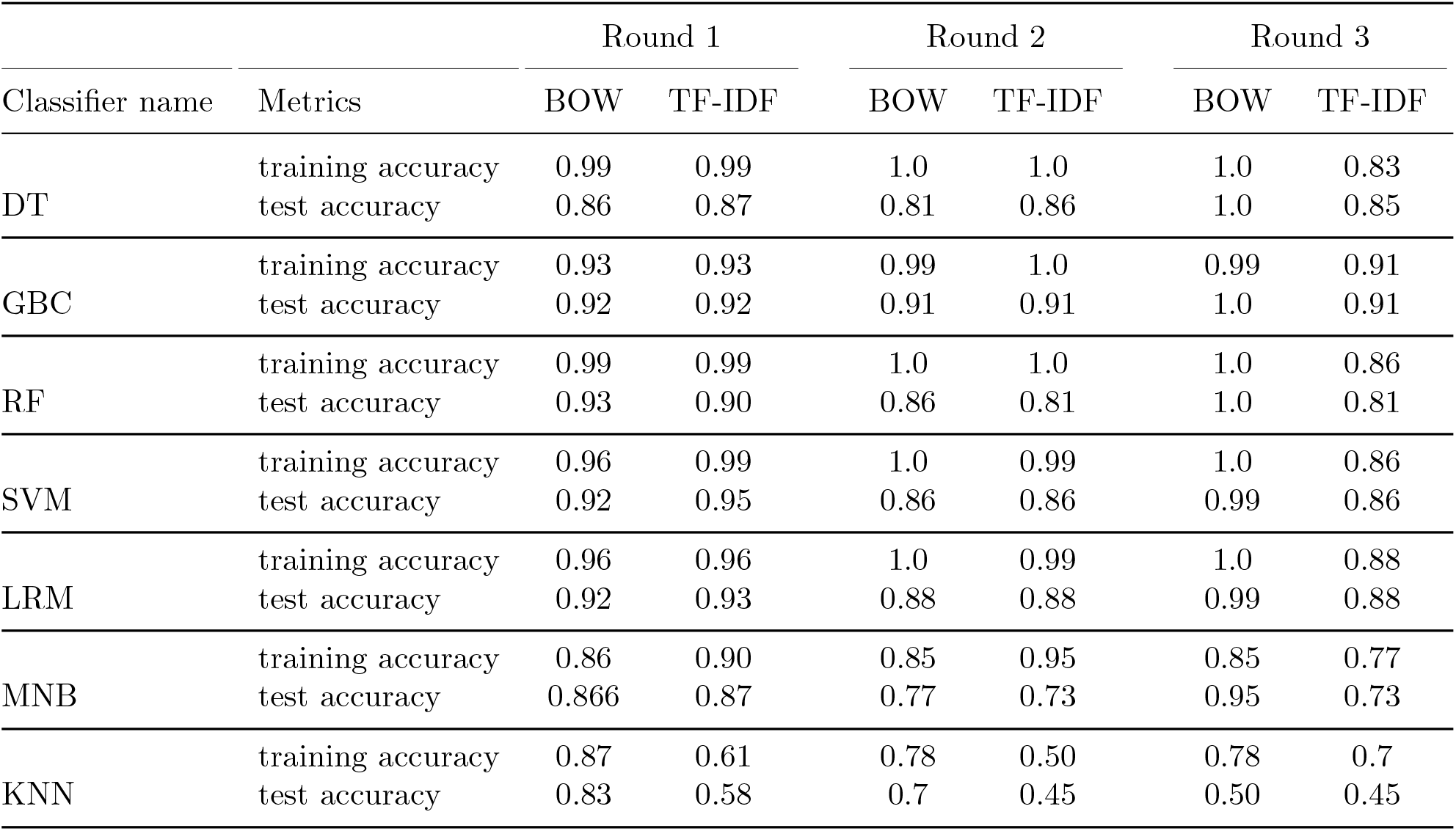
Classification accuracies using a unigram model

| Classifier name | Metrics | Round 1 |  | Round 2 |  | Round 3 |  |
| --- | --- | --- | --- | --- | --- | --- | --- |
|  |  | BOW | TF-IDF | BOW | TF-IDF | BOW | TF-IDF |
| DT | training accuracy | 0.99 | 0.99 | 1.0 | 1.0 | 1.0 | 0.83 |
|  | test accuracy | 0.86 | 0.87 | 0.81 | 0.86 | 1.0 | 0.85 |
| GBC | training accuracy | 0.93 | 0.93 | 0.99 | 1.0 | 0.99 | 0.91 |
|  | test accuracy | 0.92 | 0.92 | 0.91 | 0.91 | 1.0 | 0.91 |
| RF | training accuracy | 0.99 | 0.99 | 1.0 | 1.0 | 1.0 | 0.86 |
|  | test accuracy | 0.93 | 0.90 | 0.86 | 0.81 | 1.0 | 0.81 |
| SVM | training accuracy | 0.96 | 0.99 | 1.0 | 0.99 | 1.0 | 0.86 |
|  | test accuracy | 0.92 | 0.95 | 0.86 | 0.86 | 0.99 | 0.86 |
| LRM | training accuracy | 0.96 | 0.96 | 1.0 | 0.99 | 1.0 | 0.88 |
|  | test accuracy | 0.92 | 0.93 | 0.88 | 0.88 | 0.99 | 0.88 |
| MNB | training accuracy | 0.86 | 0.90 | 0.85 | 0.95 | 0.85 | 0.77 |
|  | test accuracy | 0.866 | 0.87 | 0.77 | 0.73 | 0.95 | 0.73 |
| KNN | training accuracy | 0.87 | 0.61 | 0.78 | 0.50 | 0.78 | 0.7 |
|  | test accuracy | 0.83 | 0.58 | 0.7 | 0.45 | 0.50 | 0.45 |

**Table 4:** The average classification accuracies of *BOW*_*n_gram*_ and *TF* − *IDF*_*n*_*_*_*gram*_

| Metrics | *unigram* |  | *n - gram* |  |
| --- | --- | --- | --- | --- |
|  | BOW | TF-IDF | BOW | TF-IDF |
| training accuracy | 0.94 | 0.97 | 0.79 | 0.89 |
| test accuracy | 0.92 | 0.81 | 0.63 | 0.59 |

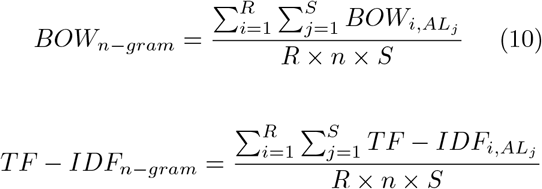

where :

- *AL*= {DT,CBC,RF,SVM,LBM,MNB,KNN},
- *S* = *Size*(*AL*),
- *n* ≥ 1, *n* ∈ ℕ,

Table 5 represents the accuracy score of applying LSTM neural network to detect news authenticity. The average training and test accuracy is computed with respect to regular word order and random shuffling of the words. The results from Table 5 demonstrate that the loss of semantics affects considerably the accuracy of news classification. This can be clearly deduced from the training accuracy, which decreased drastically from a score of 0.95 with a regular arrangement of word order to 0.53 when a random shuffling of words is performed. The test accuracy has also slightly decreased from 0.5 to 0.47, which can be explained by the small size of data used for training and testing. LSTM is a type of neural networks that is capable of learning order dependence, and the loss of semantics can clearly misclassify important information, in case such order-based neural network is considered for use.

**Table 5:** The average accuracy score of LSTM network

| Metric | Regular word order | Random word order |
| --- | --- | --- |
| training accuracy | 0.95 | 0.53 |
| test accuracy | 0.5 | 0.47 |

## 4 Conclusion

The objective of the research work presented herein consists in highlighting the importance of semantics when detecting Covid-19 related fake news. Several fake news detection approaches were discussed in literature reviews but hardly any had shed the light on the impact of semantics when applying these approaches. The proposed CNN-based color hashmap method put much emphasis on finding the correlation between semantics and the strength of a fake news classifier. The experimental results demonstrated a poor performance of the proposed classifier when semantics are ignored. Other widely used neural network models for NLP and machine learning models have also been considered to investigate the importance of word order in fake news classification. Future work will be focused on creating a CNN-based color hashmap classifier where anterior and posterior neighbors are included.

## Data Availability

https://github.com/cuilimeng/CoAID

